# The Geographical Gap in Leading Medical Journals - a Computational Audit

**DOI:** 10.1101/2022.12.12.22283368

**Authors:** Oscar Brück

## Abstract

**Background:** Auditing geographical representation in medical publishing could help to mitigate possible national and regional disparities.

**Methods:** Using the Web of Science indexing database, we collected bibliometric data of original research articles published between 2010-2019 in *The New England Journal of Medicine, Nature Medicine, Journal of the American Medical Association, The BMJ*, and *The Lancet*. We studied the corresponding authors’ geolocation in regard to publication and citation count, their temporal evolution, and the journals’ and citing organizations’ nationality.

**Results:** We identified 10,558 articles. Based on the nationality of the corresponding authors’ institutes, only 32 countries published more than 10 publications in 10 years equaling to 98.9% of all publications. English-speaking countries USA (48.2%), UK (15.9%), Canada (5.3%), and Australia (3.2%) were most represented, but with a declining trend in recent years. Normalized to their accumulated citations, 9/32 countries were associated with ≥10% publication excess, of which USA (n=1,174 publications) and UK (n=410) accounted for 85.7%. Similar findings were replicated at the municipal level where all top 10 most productive cities were located in USA (n=7), UK (n=2), or Canada (n=1), and 21 out of 25 most productive cities published more articles than predicted based on their accumulated citations. Finally, we discovered that both journals published, and researchers cited more commonly research conducted in the same country.

**Discussion:** The audit revealed Anglocentric dominance, domestic preference occurring in both journals and citation selection, and increased geographical representation in recent years in medical publishing.

## BACKGROUND

Defined as the yearly-averaged citation frequency of research articles, journal impact factors (JIF) were initially designed to rank journals by their influence^1^. JIFs have also been used for non-intended purpose. At the individual level, a successful publication track record has been interpreted as an ability to conduct high-quality research permitting both career advancements and increased chances of research funding. At the institutional level, harboring scientists with publications in journals with high JIF creates a positive feedback loop, which further attracts human and financial resources creating polarization among research centers.

Monitoring diversity and equity in academic publishing are fundamental to mitigate possible disparities. By signing the “Joint commitment for action on inclusion and diversity in publishing” more than 50 publishers representing 15,000 scientific journals will participate by collecting data on gender and ethnical representation and identify bias in the editorial and review processes^2^. However, decades of publication metadata stored in journal indexing databases such as the Web of Science (WoS), Scopus, PubMed, and Google Scholar could reveal the extent and temporal development of inequity in medical publishing.

In this retrospective cohort study, we examined possible bias related to the geographical location of the conducted research to the number of publications and citations. We also interrogated publishing and citing patterns based on the geographical origin of the conducted research.

## METHODS

### Data collection

We selected the five journals publishing mainly original articles and ranked highest in the Journal Citation Reports 2022 JIF in the field of medicine. These included *New England Journal of Medicine* (NEJM), *Nature Medicine* (NatMed), *Journal of the American Medical Association* (JAMA), *The BMJ*, and *Lancet*. We included all original articles (n=10,558) published in 2010-2019. More recent publications were not included due to unequal opportunities of being cited and to avoid bias related to COVID-19 pandemic. Using the WoS database provided by Clarivate Plc and the query “(((SO=(NATURE MEDICINE OR LANCET OR NEW ENGLAND JOURNAL OF MEDICINE OR JAMA JOURNAL OF THE AMERICAN MEDICAL ASSOCIATION OR BMJ BRITISH MEDICAL JOURNAL)) AND DT=(Article)) AND PY=(2010-2019))”, we downloaded publication metadata including author names, page length of the article, address of the corresponding author, and total number of citations. We could also query international citing patterns based on the digital object identifiers of the 10,558 original articles. National population data were downloaded from the World Bank database https://data.worldbank.org/indicator/SP.POP.TOTL, and municipal data from the world.cities dataset of the utils R package.

### Preprocessing variables of interest

We defined the impact of an article by its average citation count per year. We text-mined the number of authors, by calculating the frequency of the semicolon “;” delimiter between author names and added 1. To geolocate the primary institutes where the research has been performed, we identified the latitude and longitude coordinates of the address(es) of the corresponding author(s) with the ggmap library employing Google’s Geocoding API. For non-successful matches, we geolocated only the city and country of the address.

### Indices

We defined the international research impact (IRI) as the relationship between the total number of publications (10-fold logarithm transformed) and their cumulative citations (10-fold logarithm transformed) grouped by the research institute of the corresponding authors. In this study, IRI was used separately when comparing countries and cities.

We defined the Domestic Self-Citation Index (DSCI) as:

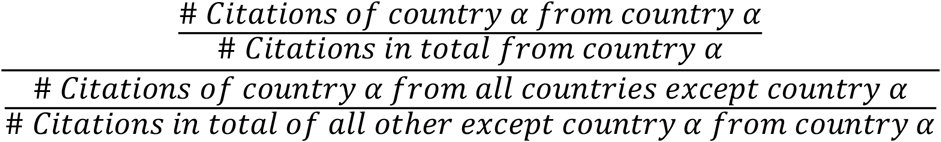

where “# Citations” represents the number of citations and α any country of interest.

### Statistical analysis

We performed comparisons of two continuous variables with the Wilcoxon rank-sum test (unpaired, two-tailed) and three or more continuous variables with the Kruskal-Wallis test. For categorical variables, we used the χ^2^ test. We adjusted p values with the Benjamini–Hochberg correction. We performed hierarchical clustering with Euclidean distance metrics and the Ward.d2 method. To compare two linear regression slopes, we tested the T-test significance of the interaction term. We conducted statistical analyses and visualizations with R 3.5.1. using base, tidyverse, fastDummies, maps, reshape2, ggmap, data.table, countrycode, ggpubr, ggrepel, rstatix, ggdendro and dendextend libraries.

## RESULTS

### National publication productivity does not imply high citation frequency

Of 10,558 original articles published between 2010-2019 in *NEJM* (n=2,966; 28.1%), *JAMA* (n=1,833; 17.4%), *NatMed* (n=1,577; 14.9%), *Lancet* (n=2,462; 23.3%), and *BMJ* (n=1,720; 16.3%), corresponding authors were affiliated to institutes from 77 countries. Each article included 1.2 mean affiliations [range 1-15] equaling to 10,732 total unique entries, of which 100.0% (n=10,730) could be geolocated. When focusing on countries with ≥10 original articles in 10 years in any of these five medical journals, only 32 countries were included representing 98.9% (n=10,613) affiliations highlighting the geographical exclusivity of medical research.

Of these, English-speaking countries were overrepresented with almost 3/4 of publications from the United States of America (USA, 48.2%), the United Kingdom (UK, 15.9%), Canada (5.3%), and Australia (3.2%, Table 1). National population-normalized publication frequencies correlated with total publication number (corr 0.69 p<0.001, Spearman test). Following population-normalization, the most represented countries included Denmark (31.1 articles per million people), UK (25.1), and Switzerland (24.8, Table 1).

**Table 1.** Publication metrics by country.

| Country | # Citations | # Citations/<br>Year | # Articles | # Population | # Citations/<br>Article/<br>Population/<br>Year | # Articles/<br>Population | # Excess<br>Articles | % Excess<br>Articles |
| --- | --- | --- | --- | --- | --- | --- | --- | --- |
| Australia | 20443.2 | 28.9 | 339 | 25.7 | 1.1 | 13.2 | -22.1 | -6.5 |
| Austria | 1594.3 | 29.8 | 36 | 9.0 | 3.3 | 4.0 | -1.9 | -5.2 |
| Bangladesh | 162.4 | 14.6 | 11 | 166.3 | 0.1 | 0.1 | 6.0 | 54.3 |
| Belgium | 5111.6 | 40.2 | 82 | 11.6 | 3.5 | 7.1 | -24.1 | -29.4 |
| Brazil | 3094.9 | 37.1 | 42 | 214.0 | 0.2 | 0.2 | -26.1 | -62.1 |
| Canada | 30632.0 | 28.4 | 560 | 38.2 | 0.7 | 14.6 | 43.8 | 7.8 |
| China | 15919.5 | 39.6 | 225 | 1412.4 | 0.0 | 0.2 | -64.5 | -28.7 |
| Denmark | 6633.5 | 21.2 | 182 | 5.9 | 3.6 | 31.1 | 48.5 | 26.6 |
| Finland | 1679.8 | 26.1 | 40 | 5.5 | 4.7 | 7.2 | 0.3 | 0.8 |
| France | 24133.5 | 33.7 | 386 | 67.5 | 0.5 | 5.7 | -32.1 | -8.3 |
| Germany | 21318.0 | 36.0 | 376 | 83.1 | 0.4 | 4.5 | 1.3 | 0.3 |
| Greece | 733.3 | 31.9 | 14 | 10.7 | 3.0 | 1.3 | -5.1 | -36.2 |
| India | 1278.1 | 21.3 | 34 | 1393.4 | 0.0 | 0.0 | 2.8 | 8.4 |
| Ireland | 722.1 | 29.5 | 17 | 5.0 | 5.9 | 3.4 | -1.8 | -10.7 |
| Israel | 2047.4 | 22.0 | 47 | 9.4 | 2.3 | 5.0 | -0.3 | -0.5 |
| Italy | 6699.9 | 35.3 | 133 | 59.1 | 0.6 | 2.3 | -1.7 | -1.3 |
| Japan | 5982.8 | 26.9 | 112 | 125.7 | 0.2 | 0.9 | -9.9 | -8.8 |
| Kenya | 453.4 | 23.7 | 17 | 55.0 | 0.4 | 0.3 | 4.5 | 26.7 |
| Netherlands | 17730.4 | 25.9 | 374 | 17.5 | 1.5 | 21.3 | 55.6 | 14.9 |
| New Zealand | 2240.5 | 27.8 | 53 | 5.1 | 5.4 | 10.3 | 1.8 | 3.4 |
| Norway | 1935.5 | 19.6 | 62 | 5.4 | 3.6 | 11.5 | 17.0 | 27.5 |
| Pakistan | 377.4 | 11.6 | 13 | 225.2 | 0.1 | 0.1 | 2.4 | 18.5 |
| Saudi Arabia | 638.4 | 49.4 | 12 | 35.3 | 1.4 | 0.3 | -4.9 | -40.6 |
| Singapore | 1173.0 | 23.2 | 23 | 5.5 | 4.3 | 4.2 | -5.9 | -25.6 |
| South Africa | 2368.6 | 25.5 | 58 | 60.0 | 0.4 | 1.0 | 4.3 | 7.3 |
| South Korea | 1656.9 | 39.1 | 34 | 51.7 | 0.8 | 0.7 | -5.2 | -15.3 |
| Spain | 8909.0 | 39.6 | 97 | 47.3 | 0.8 | 2.0 | -76.3 | -78.7 |
| Sweden | 7303.3 | 21.2 | 199 | 10.4 | 2.0 | 19.1 | 53.6 | 26.9 |
| Switzerland | 10218.5 | 29.9 | 216 | 8.7 | 3.4 | 24.8 | 20.4 | 9.4 |
| Thailand | 946.8 | 26.7 | 17 | 70.0 | 0.4 | 0.2 | -6.9 | -40.6 |
| UK | 85627.5 | 24.4 | 1691 | 67.3 | 0.4 | 25.1 | 410.5 | 24.3 |
| USA | 305130.0 | 28.0 | 5111 | 331.9 | 0.1 | 15.4 | 1174.5 | 23.0 |

When examining the international research impact (IRI) by countries, publication count explained 96.5% of total citations corresponding to an excellent statistical correspondence (Fig. 1A). However, we also observed unexpected discrepancies. We identified 19/32 countries with over 10% publication excess or deficit when normalized by their citation frequencies. Based on the relative number of articles, Spain published 78.7% (n=76) and Brazil 62.1% (n=26) fewer and Bangladesh 54.3% (n=6) too many articles than expected. Based on the absolute number of articles, 1,174 (23.0%) and 410 (24.3%) excess articles were published by corresponding authors affiliated to an organization located in the USA and UK, respectively. Together, USA and UK accounted for 85.7% of all articles published in excess (n=1,847).

**Figure 1.**
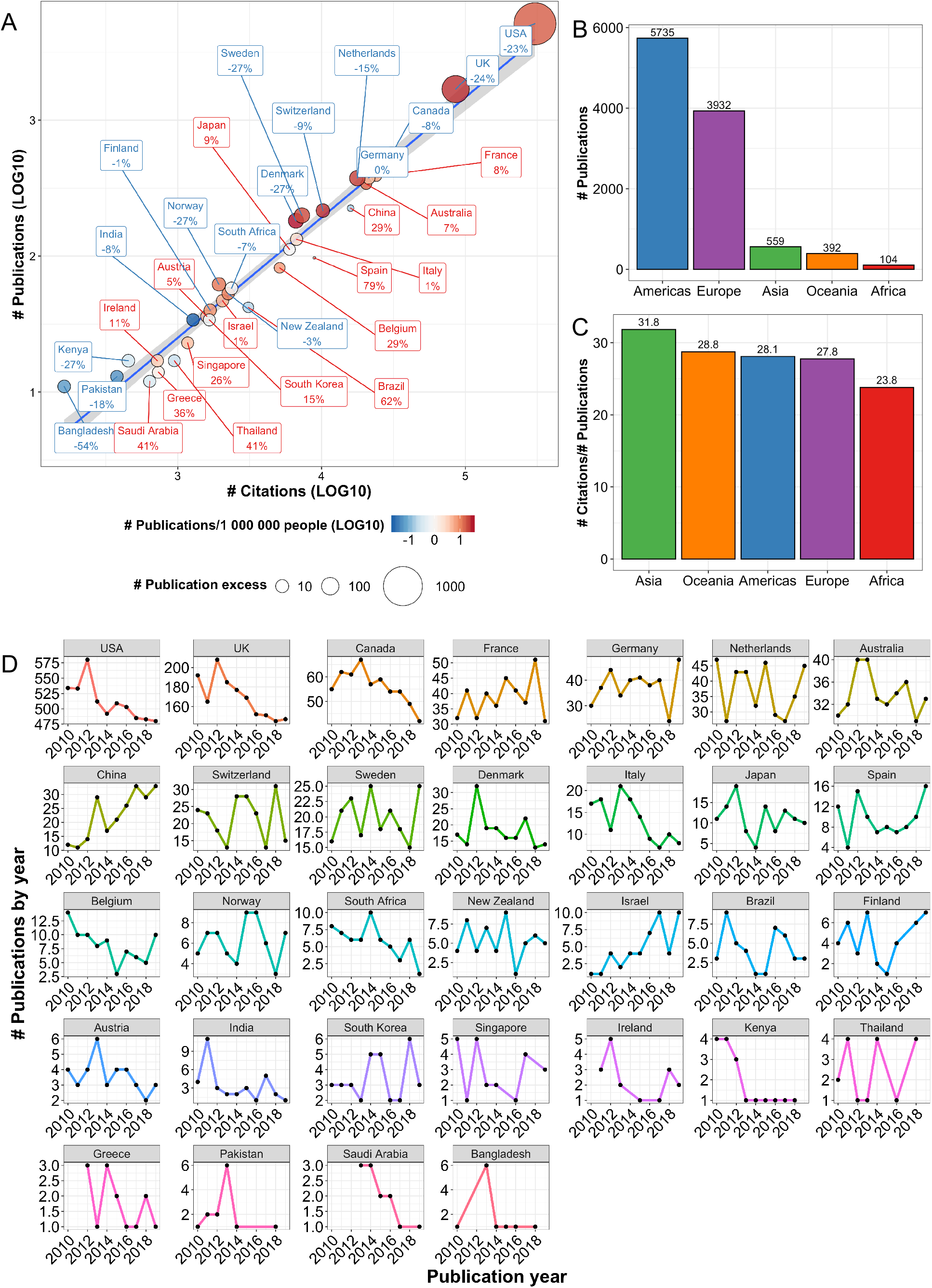
Medical publishing at the national level. (A) Linear regression of the number of citations (LOG10-transformed) and publications (LOG10-transformed) of the top 32 most productive countries. The percentage and font color indicate the proportion of excess (positive number and red font) or deficient (negative number and blue font) publications compared to the predicted number based on the citation/publication count. (B) Bar plots illustrating the number of publications and (C) citations per publication by continent. (D) Line plot illustrating the number of publications by their publishing year from the top 32 most productive countries.

Publications with a corresponding author from Saudi Arabia were associated with highest median yearly citations (49.4) per publication given their emphasis on the documentation of the Middle East Respiratory Syndrome (MERS, 7 out of 11 total publications from Saudi Arabia). However, even without MERS-related articles, the median yearly citation of publications from Saudi Arabia was the highest 41.8. Other countries with most elevated median yearly citation per publication included Belgium (40.2), and China (39.6). Lowest median citations were associated with publications geolocated in South Asia (Pakistan 11.6, Bangladesh 14.6, India 21.3) and Scandinavia (Norway 19.7, Denmark 21.2, Sweden 21.3).

When extrapolating to continental level, publication exclusivity was even more pronounced, with 89.5% of all articles originating either from the Americas (53.4%) or Europe (36.1%, Fig. 1B). However, the 5.2% publications originating from Asian countries were associated with 15.6% higher citation frequencies compared to median (27.5) and other continents (Fig. 1B-C).

We then studied how the number of publications per country has evolved during the study time period. By measuring the slope of a fitted linear regression, yearly publishing productivity in the studied journals decreased steadily in the USA (8.0 fewer articles per year), UK (5.7), Canada (1.7), and Italy (1.2, Fig. 1D). However, the number of publications originating from China (2.5) and Israel (0.89) rose, but with a lower inclination. The IRI patterns varied also temporally (Supplementary Fig. 1). Countries with numerous publications (USA, UK, Germany, Australia) gradually stepped up their yearly citation frequencies in line with rising medical journal impact factors. The average citations decreased for some countries for limited periods (France, Netherlands, China) before resuming their original increasing inclination. Heterogenous citation patterns were found in countries with 1-10 yearly publications in average.

### Publications from elite research institutes tend to accumulate fewer citations per article

The international publication selection bias was even more pronounced when studying the regional location of the corresponding authors’ institutes (Fig. 2A). The most distinct publication hot spots were situated in Northeastern USA, Central Europe, and the UK with additional dense centers in Southwestern USA and East Asia. On the contrary, the number of citations per article did not replicate similar geographical patterns (Fig. 2A). The 10 most represented cities in terms of publication number covered 36.5% of all publications and were all located in the Anglosphere: USA (n=6), UK (n=3) and Canada (n=1). The three cities with over 300 publications in 2010-2019 were Boston, USA (n=1,209, 12.2% of all publications), London, UK (n=648, 6.5%) and New York, USA (n=378, 3.8%).

**Figure 2.**
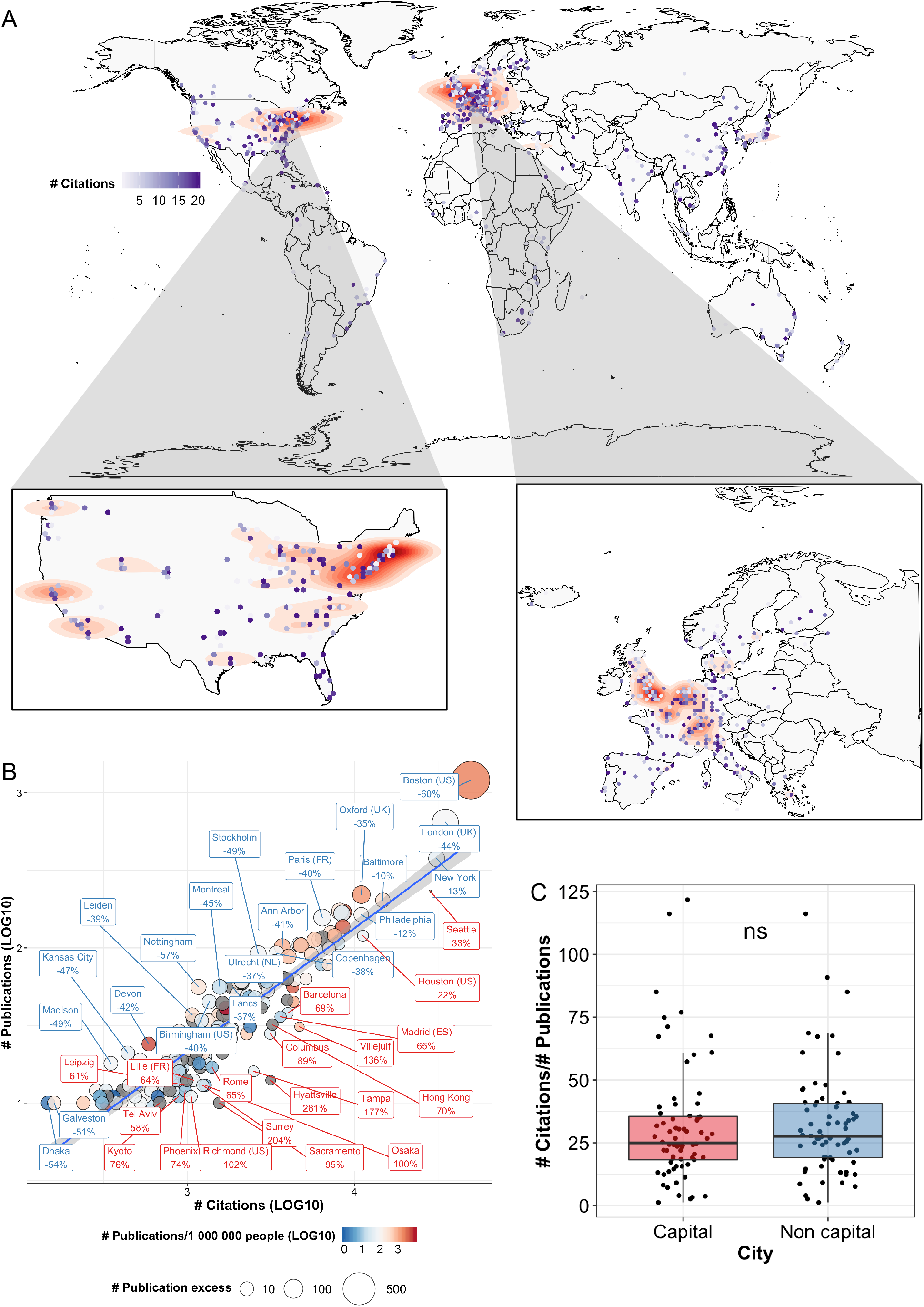
Medical publishing at the municipal level. (A) A world map with density contours indicating hot spots based on the number of publications. Blue-colored points reflect the average number of citations in that area. (B) Linear regression of the number of citations (LOG10-transformed) and publications (LOG10-transformed) by cities. The percentage and font color indicate the proportion of excess (positive number and country name in red) or deficient number of publications compared to the predicted number based on the citation/publication count. Country name abbreviations are added for city names, which are found in more than one country. (C) Box plot of the number of citations per publication in capital vs. non-capital cities (Paired Wilcoxon test).

Next, we examined municipal IRIs by including only cities with ≥10 publications in 10 years retaining 94.0% of the number of publications (n=9,327). Publication number explained 83.6% of the variability in total accumulated citations (Fig. 2B). Multiple metropolitan cities known for their established research institutes were overrepresented in terms of publication frequencies. Unexpectedly, 21 out of the 25 most productive cities accumulated less citations than predicted with linear regression suggesting selection bias. Cities with the highest absolute publication excess included Boston, USA (n=728 excess articles), London, UK (n=285) and Oxford, UK (n=76). On the contrary, cities with the highest publication deficit included Seattle, USA (n=76), Houston, USA (n=27) and Los Angeles, USA (n=6).

While the publication count decreased in the USA and UK during the 10-year follow-up, similar patterns were not as evident when examining the 25 most productive institutes (Supplementary Fig. 3A). The publishing dynamics varied with some cities publishing gradually more (New York, Stanford, Philadelphia), some less (London, Baltimore, Ann Arbor), but for most the direction was inconclusive. As in the national level, the average yearly citations-per-article increased also at the institute level and the slope tendency varied more in cities with fewer publications (Supplementary Fig. 3B). Houston stood out with higher citations/publication across follow-up, whereas publications originating from Boston, Toronto, and Copenhagen accumulated fewer citations compared to other institutes.

**Figure 3.**
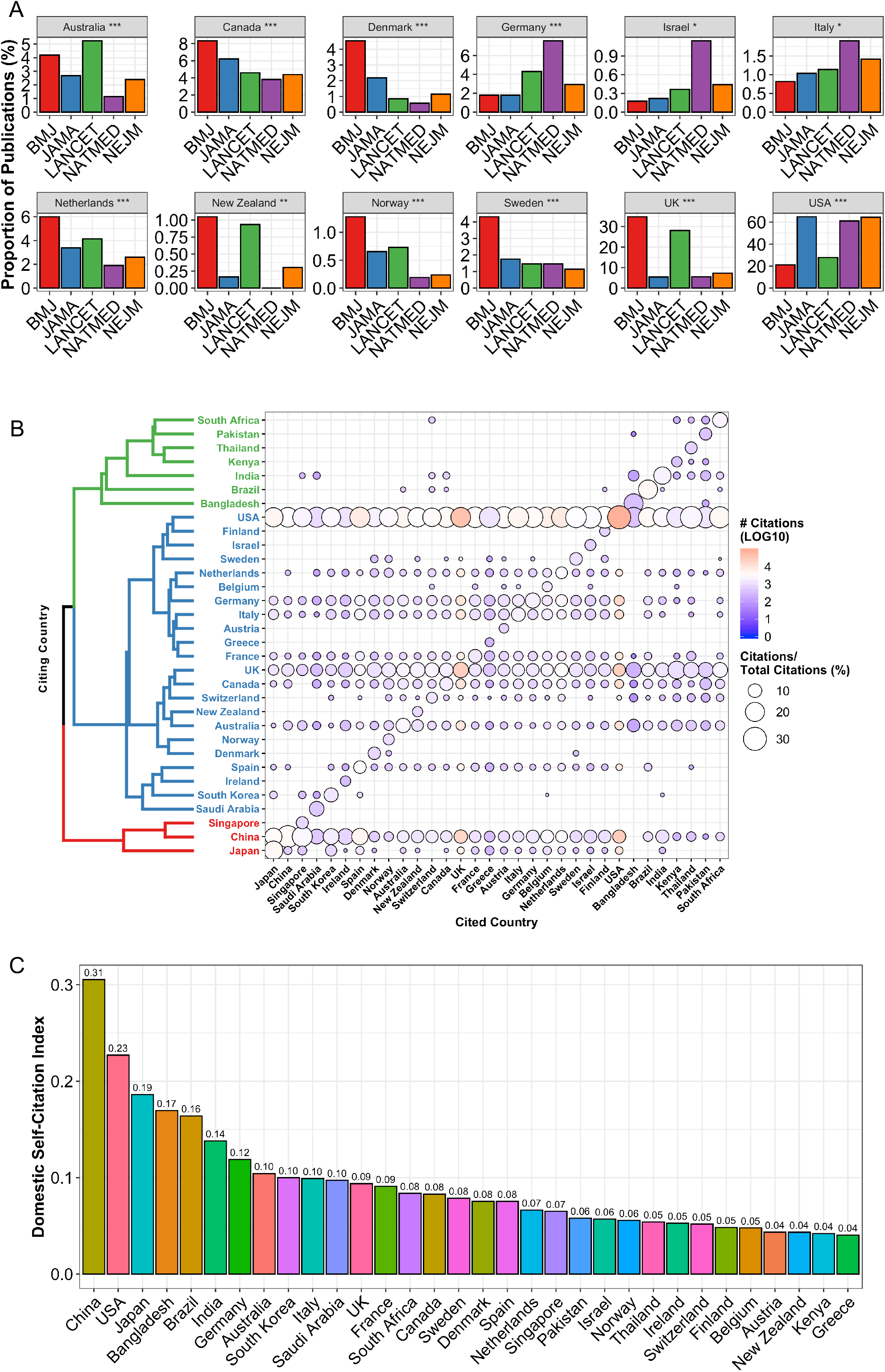
National journal and citation fingerprints. (A) Bar plot illustrating the proportion of publications in medical journals originating from different countries. Only significant results of the Kruskal-Wallis analysis are plotted. (B) Balloon plot illustrating the international citation patterns of articles published in leading medical journals. The balloon size reflects the proportion which countries have cited the most any given country (column-wise). The balloon color reflects the absolute number of citations any given country has received. Citing countries have been grouped by hierarchical clustering (Euclidean distance). (C) Bar plot illustrating the same data as in the previous plot but proportioned to how much other countries have cited any given country in relation to how much they have cited all others countries. See Methods for the definition of the Domestic Self-Citation Index.

The absolute number of publications correlated moderately with population-normalized publication frequencies (corr 0.41, p<0.001). However, the residuals of the linear regression model between publication number and citations increased (corr 0.37, p<0.001) by publication number signifying that articles originating from established research organizations tended to accumulate fewer citations than statistically would be expected. Finally, institutes from capital cities did not accumulate more citations per article compared to non-capital cities indicating that the governmental status or population of the city are poor predictors of IRI (Fig. 2C and Supplementary Fig. 2).

### Institute nationality affects both journalistic and citation patterns

Given overlap in the scientific scope of the studied five medical journals, we reasoned that the geographical coverage of publications should not differ. However, we observed substantial preponderance of publications from English-speaking countries (UK, New Zealand and Australia) in both UK-based *BMJ* and *Lancet* journals (Fig. 3A). Moreover, publications from Scandinavian countries (Norway, Sweden and Denmark), Netherlands and Canada were overrepresented in *BMJ*. Publications from Germany, Israel, and Italy were most common in *NatMed*, which is part of the British-German Springer publishing group. Publications from institutes located in the USA were most represented in *NatMed* and US-based journals *JAMA* and *NEJM*.

To conclude, we audited national citation patterns. First, countries with most publications were the most active to cite as expected (Fig. 3B, horizontal lines of recurring balloons). USA was the most common citing country in 30/32 of the most frequently publishing countries. Articles from China and Singapore were cited more commonly in publications with a corresponding author based in China.

Secondly, countries tended to show domestic preference when citing (Fig. 3B, diagonal balloons). We observed highest absolute domestic self-citation (#domestic citations/#all citations) among researchers affiliated to an institute in USA, UK, or South Africa (Supplementary Fig. 4). To highlight the proportion of self-citations compared to the proportion of citations originating from abroad, we introduce the domestic self-citation index (DSCI, see Methods). The highest DSCI associated with China, USA, and Japan, and the lowest with Austria, New Zealand, Kenya, and Greece (Fig. 3C). National DSCI correlated with their total accumulated citation count (corr 0.70, p<0.001) and publication count (corr 0.46, p=0.015), but not with the absolute count (corr 0.032, p=ns) nor proportion of excess articles (corr -0.060, p=ns). Domestic self-citation accounted for 74.5% of total national citation count (corr 0.86, p<0.001) indicating the serious bias related to citation count for evaluating manuscript impact.

Lastly, we observed that national citation patterns formed distinct clusters (Fig. 3B, horizontal colored groups). The largest cluster was composed of the Anglosphere, European countries, South Korea, and Saudi Arabia. The second largest cluster included developing countries whereas China, Japan, and Singapore formed the third cluster. In summary, similar geographical location, development index and health challenges account for citation trends at the national level.

## DISCUSSION

Available bibliometric data can reveal important information on the equity and representation in medical research. Longitudinal data permits the study of temporal trends and can be repurposed for monitoring of diversity. Here, we presented geographical publication disparity in five leading medical journals based on JIFs between 2010-2019.

We limited the scope of this study to available geographical data recorded in scientific indexing databases. Therefore, for example ethnical, career-stage, and gender disparity was not examined. Similarly, the number of submissions and geographical coverage of reviewers are unavailable, while these could help to further interpret the results.

First, we described the overrepresentation of distinct countries and research organizations. Almost 3/4 of all publications with a corresponding author originated from an institute in USA, UK, Canada, or Australia indicating medical publishing is Anglocentric. Similar findings have been reported both in leading medical journals between 1971-2005 and in the field of pathology between 2000-2006 suggesting both an interdisciplinary and long-lasting phenomenon^3,4^. We showed that the number of articles from institutes in USA, UK and Canada was decreasing, while publications from China and Israel were more frequent reflecting a gradual transition in international representation.

Second, by comparing IRIs our analysis highlighted an undocumented discrepancy favoring notably highly productive institutes and countries. For instance, 86% of all articles published in excess based on their IRI originated from USA or UK. At the municipal level, 21/25 of the leading centers were found to publish more articles than expected. In particular, the number of publications with a corresponding author located in Boston exceeded 60% its predicted quantity.

Multiple factors could be involved. For instance, previous studies have indicated a pronounced correlation between national gross domestic product and publishing in top medical journals^5,6^. The publication fees require considerable research funding, which are primarily available in high-income countries making scientific publishing an unintended luxury. Moreover, our data indicated higher proportions of articles from UK, Australia, Canada and New Zealand in UK-based journals (*BMJ* and *Lancet*), while corresponding authors from USA were overrepresented in American journals (*JAMA* and *NEJM*). UK-affiliated research published in UK-based journals (*BMJ*, 28.1%; Lancet, 34.8%; mean 30.8%) was almost 5-fold compared to the average proportion in *NEJM, NatMed* and *JAMA* (6.3%). Importantly, journals such as *Lancet* advice authors to consider diversity when inviting coauthors^7^. Yet, further comparative studies on submission rates are required to better understand the reasons and to exclude domestic preference at review or editorial level, which could undermine public trust in scientific publishing and possibly found new journals from underrepresented countries. Importantly, our data indicated that less-publishing countries and institutes tended to accumulate higher citations, which could reflect both interest and ultimately elevate further JIFs. Diversity and inclusion of developing countries would benefit the scientific community as demonstrated during the SARS, MERS, and COVID-19 epidemics, facilitate the adoption of health policies globally, and diversify medical research by increasing international collaborations^8^.

When examining international citation patterns, we identified considerable domestic preference. The nationality of the research institute accounted for 74.5% of the international citation variance. In addition, the DSCI varied 8-fold between the studied countries emphasizing that further research in citation patterns is needed. The results are in line with a study reporting that 85% of the variability of the scientific journalism coverage of 22 newspapers focused on domestic research^9^. However, we also observed international citing clusters reflecting likely similar research interests and collaborations.

In summary, this computational audit of author affiliations in top medical journals revealed the Anglocentric dominance embodied as publication excess in relation to their accumulated citation and domestic preference by journals. Based on longitudinal data, the international representation gap in top medical journals is gradually decreasing.

## Data Availability

Codes and a 100-row data example are available at https://github.com/obruck/International-Research-Impact. Raw data can be downloaded from Clarivate Web of Science, with instructions provided in the Github repository.

## ADDITIONAL INFORMATION

## ACKNOWLEDGMENTS

The author wishes to thank Susanna Lallukka-Brück for her patience, understanding, and insightful comments. The author is grateful for Olli Dufva and to the members of the Hematoscope Lab for discussion and comments. Certain data included herein are derived from Clarivate Web of Science. © Copyright Clarivate 2022. All rights reserved.

## AUTHORS’ CONTRIBUTIONS

Conception and design; Collection and assembly of data; Data analysis; Manuscript writing; Manuscript editing; Data interpretation; Final approval of manuscript: O.B.

## ETHICS APPROVAL AND CONSENT TO PARTICIPATE

The study complied with the Declaration of Helsinki.

## COMPETING INTERESTS

O.B. declares no Competing Non-Financial Interests but the following Competing Financial Interests: consultancy fees from Novartis, Sanofi, and Amgen, outside the submitted work.

## FUNDING INFORMATION

This study was supported by research grants from the Helsinki University Hospital.

## Notes

### Competing Interest Statement

O.B. declares the following Competing Financial Interests: consultancy fees from Novartis, Sanofi, and Amgen, outside the submitted work.

