## Supplementary Figures for "The Geographical Gap in Leading Medical Journals - a Computational Audit"

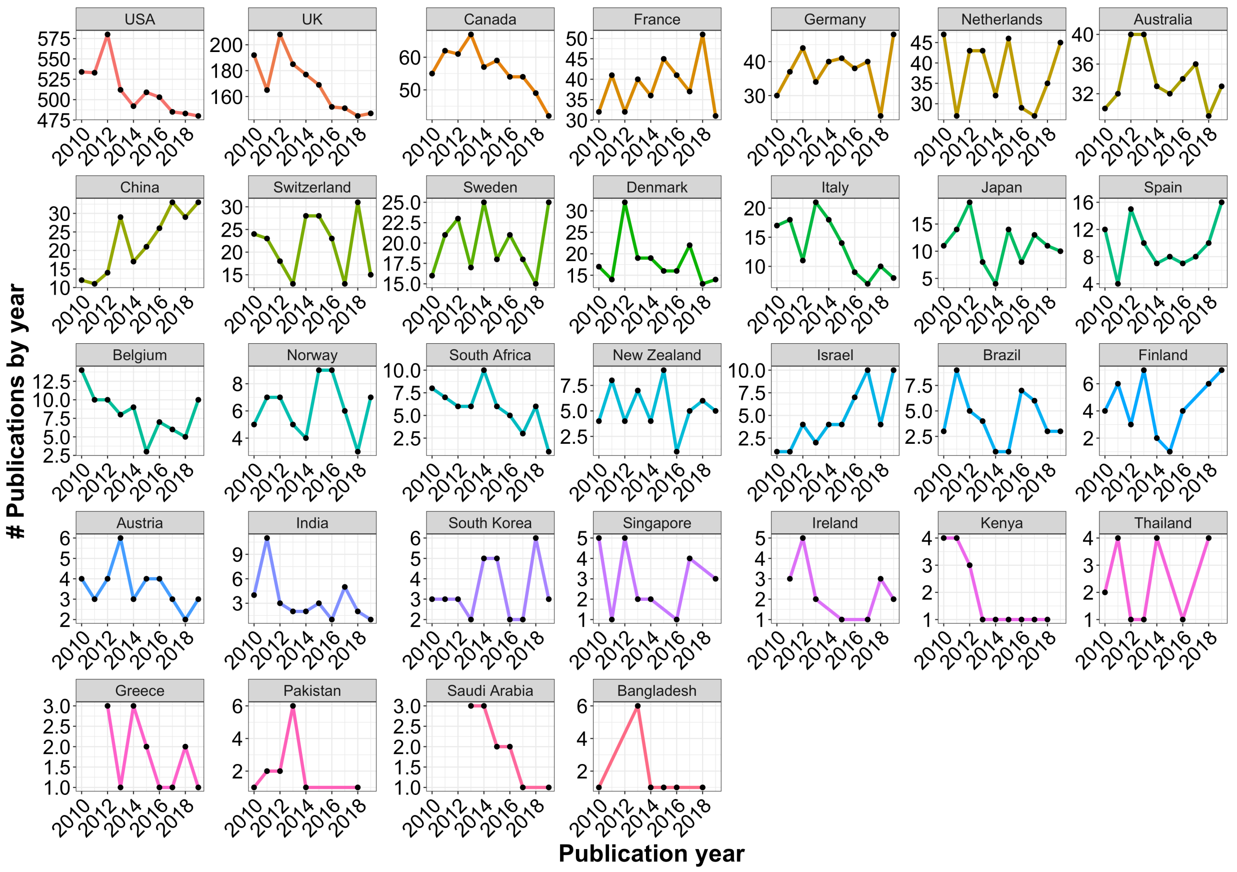


Supplementary Figure 1. Line plot illustrating the number of citations per publication by their publishing year for the top 32 most productive countries.

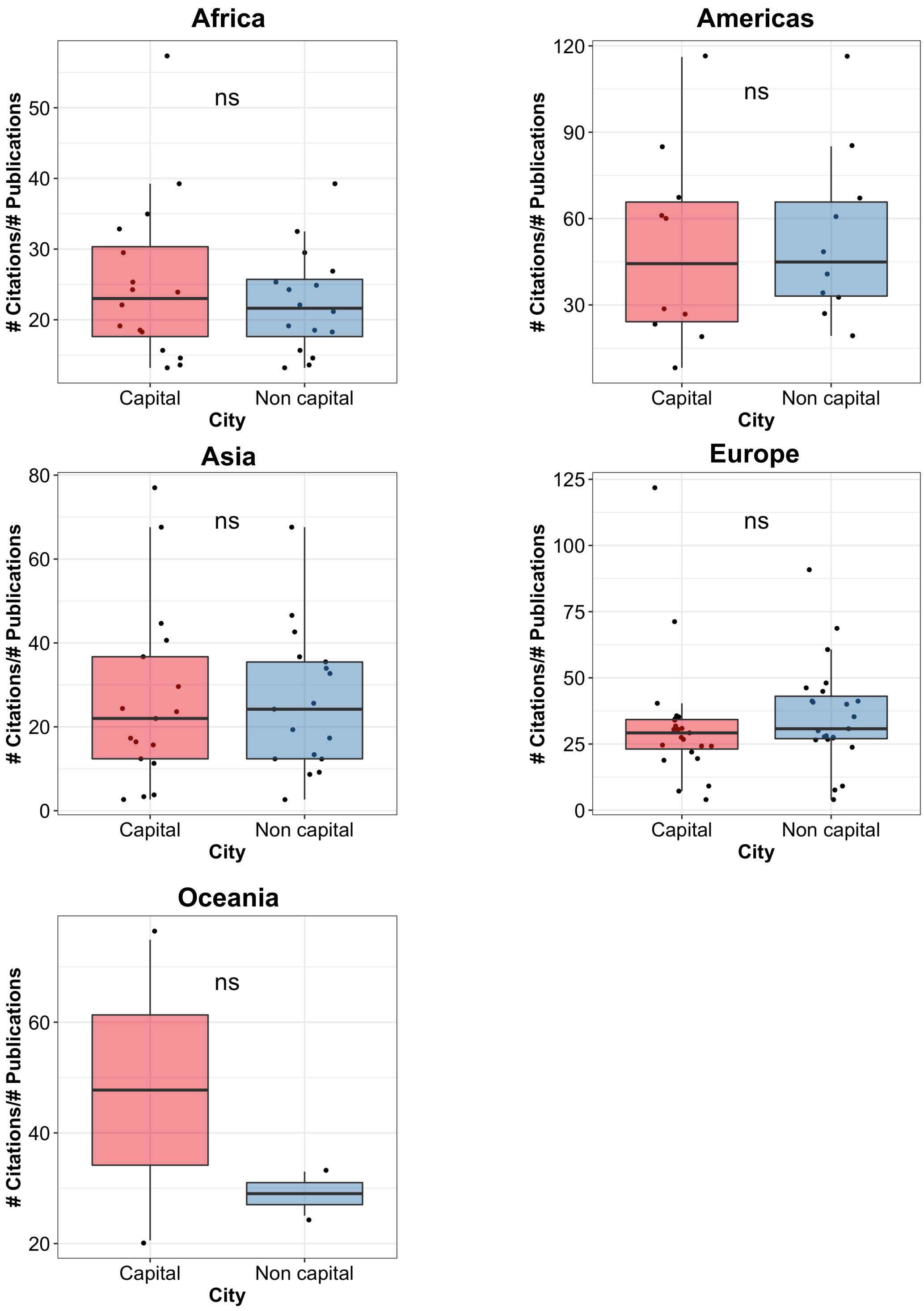


Supplementary Figure 2. Box plots illustrating the number of citations per publication in capital vs. non-capital cities grouped by continents (Paired Wilcoxon test).

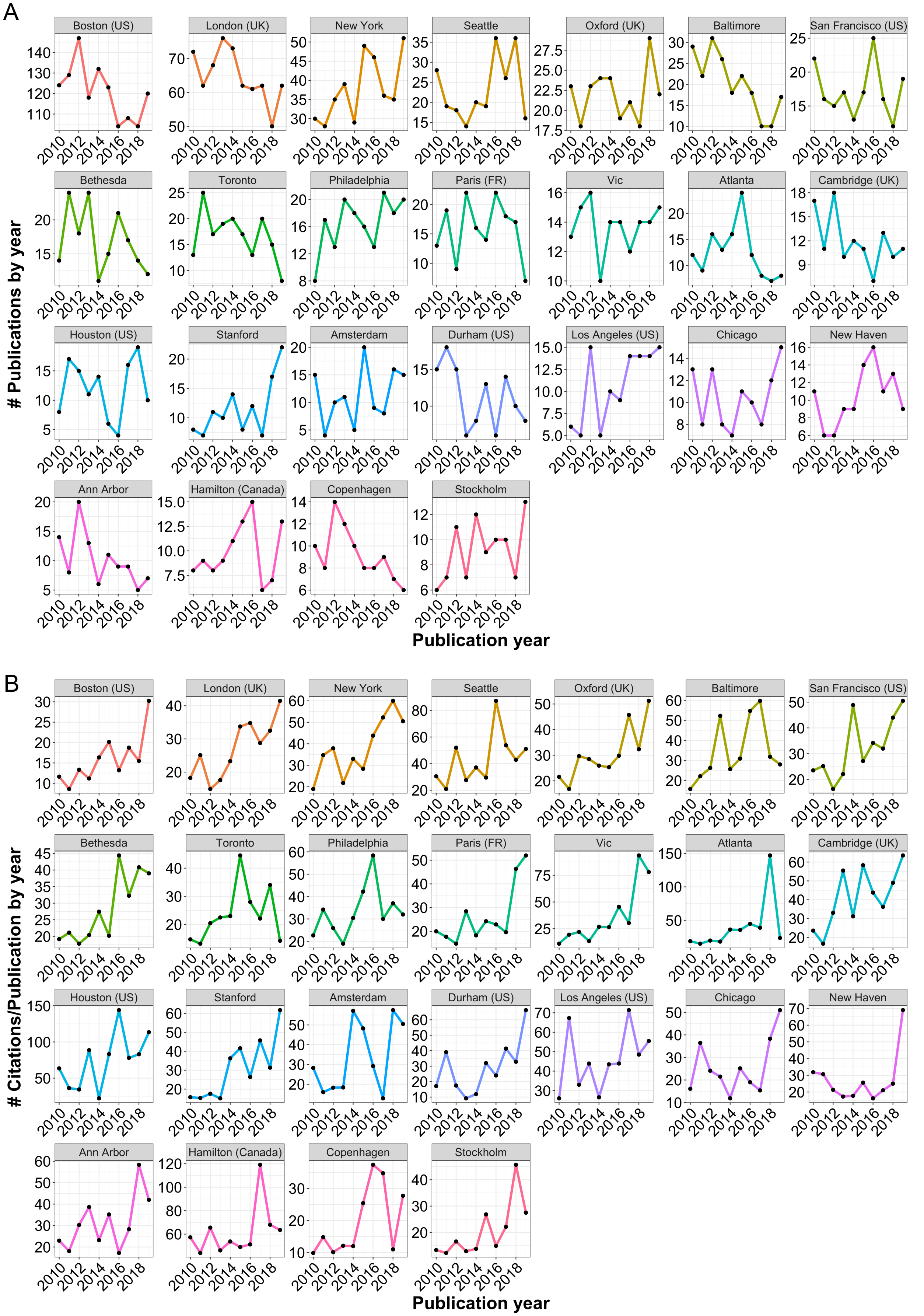


Supplementary Figure 3. Line plot illustrating the number of (A) publications and (B) yearly-averaged citations per publication by their publishing year for the top 25 most productive cities.

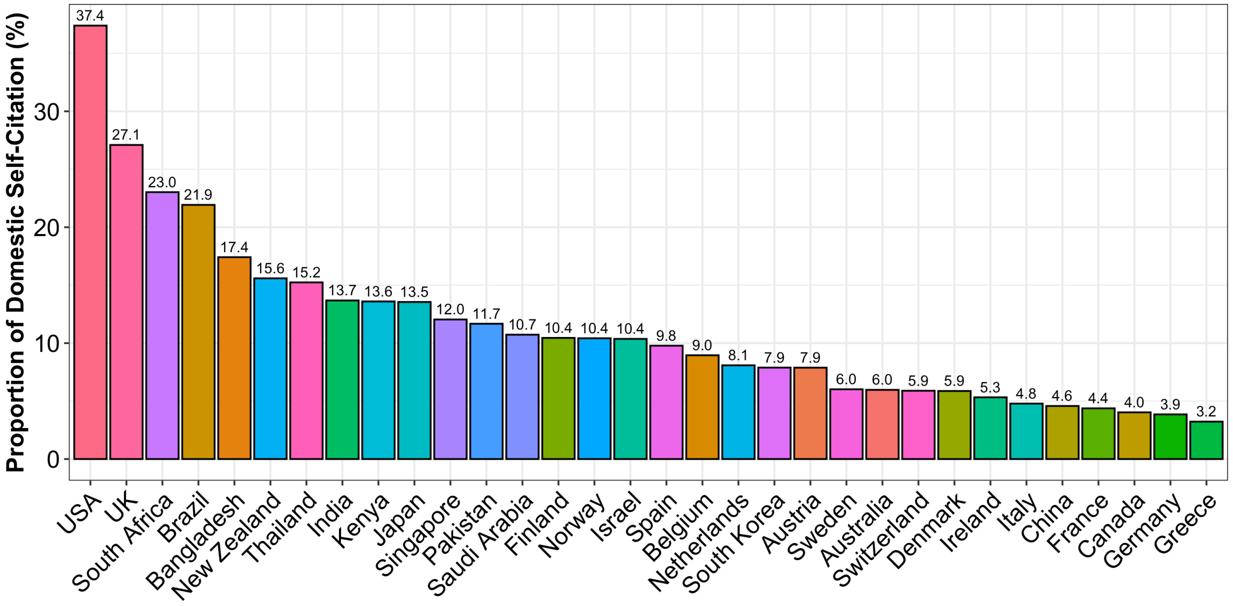


Supplementary Figure 4. Bar plot illustrating the proportion of domestic citations of all (domestic + international) citations by citing country.
